# A Multi-Agent Large Language Model Reasoning Engine for Early Detection of Pediatric Growth Disorders

**DOI:** 10.64898/2026.08.28.26361655

**Authors:** Naveed Rabbani, Jannik Mettner, Kyungjoon Lee, Carmen L. Soto-Rivera, Alexander Windberger, Karl Santiago, Jonathan Hatoun, Emily Trudell Correa, Louis Vernacchio, Isaac Kohane

## Abstract

Routine childhood growth surveillance is a cornerstone of pediatric care. Growth pattern abnormalities are often early manifestations of chronic disease. Yet subtle abnormalities are frequently underrecognized, leading to diagnostic delays and avoidable morbidity. We introduce **sprout** (**S**ystem for **P**ediatric **R**ecognition **O**f **U**ndiagnosed **T**rajectories), a generalized, multi-agent large language model (LLM) reasoning system designed to identify a broad spectrum of pediatric growth-related conditions from longitudinal electronic health records (EHRs) earlier than standard clinical practice.

Using a large pediatric primary care EHR dataset, we developed and validated **sprout** as a two-stage system. First, a highly specific LLM screener flags concerning longitudinal growth patterns. Second, an Orchestrator module coordinates a multidisciplinary panel of LLM agents to generate a ranked differential diagnosis. To correct systemic reasoning errors, a Trainer module injects meta-knowledge into the panel via a dedicated “Learner” agent. Diagnostic capability was evaluated using a walk-forward, visit-by-visit simulation leading up to the diagnosis date.

The **sprout** screener model achieved 98% (83/85) specificity and 28% (9/32) sensitivity on a gold-standard dataset of pediatric primary care patients when evaluated one year before the index date, and 100% specificity and 47% sensitivity when evaluated using longitudinal data up to the day of diagnosis. When applied to 300 control patients (i.e., healthy or undiagnosed), the screener flagged 15. Subsequent expert panel review confirmed high suspicion for undiagnosed pathology in 33% (5/15) of these cases. In chronological walk-forward validation on disease cases, the diagnostic engine identified conditions well before standard-of-care documentation. One year prior to clinical diagnosis, the system achieved sensitivities of 81% for type 1 diabetes mellitus, 56% for pituitary disorders, and 44% for celiac disease.

The **sprout** multi-agent system demonstrates the ability to detect a significant portion of latent growth-related pediatric conditions months to years before current clinical standards while minimizing false positives. These results support its potential as a decision support tool for reducing diagnostic delays in pediatric care.

## 1. Introduction

The assessment of growth is an essential component of preventive pediatric care.^1^ Growth pattern abnormalities are often early manifestations of chronic disease, such as inflammatory bowel disease, type 1 diabetes mellitus, and others. Despite their importance, however, misinterpretation and diagnostic errors of growth are common,^2,3^ leading to delays in diagnosis and avoidable morbidity. Take, for example, inflammatory bowel disease, which affects an estimated 100,000 children in the United States.^4^ Despite its prevalence, diagnostic delays on the order of several months are the norm for children with inflammatory bowel disease such as ulcerative colitis or Crohn’s disease.^5^ Similarly, Turner’s syndrome, despite being a genetic condition present from birth, has a median age of diagnosis of 15 years.^6^

Prior studies have attempted to address this gap through two mechanisms. Early work by Kohane *et al*. proposed a piecewise regression-fitting method for detecting abnormal growth trends as a means for identifying a wide range of growth-related conditions in children.^7,8^ Recent attempts have developed disease-specific machine learning models run against medical records to close the gap in diagnosis for such conditions, alerting clinicians when the probability of disease exceeds a prespecified threshold. Examples of the latter include work by Daniel *et al*. who developed a SuperLearner model capable of diagnosing type 1 diabetes mellitus with a sensitivity of 71.6% when applied to patient records 90 days before documented diagnosis had otherwise been made.^9^ Similarly, Dreyfuss *et al*. developed an XGBoost model capable of diagnosing celiac disease seropositivity 1 year earlier with an AUC of 0.86.^10^

With advances in large language models (LLMs), which are now capable of performing clinical reasoning tasks,^11^ there is an opportunity to develop a more generalized clinical reasoning engine capable of addressing this problem. This is both useful and necessary in a general pediatrics practice where individual chronic disorders are infrequent but in aggregate are frequent. In this study we describe a multi-agent large-language-model-based clinical reasoning system to narrow the diagnostic delay in growth-related chronic conditions of childhood. This system, which we call SPROUT: **S**ystem for **P**ediatric **R**ecognition **O**f **U**ndiagnosed **T**rajectories, was developed and validated on a large dataset of clinical records from a pediatric primary care health system.

## 2. Methods

We present a retrospective validation of SPROUT, a two-part LLM-based clinical reasoning system for the early identification of growth-related conditions in children. The study was a collaboration between Harvard Medical School and the Pediatric Physicians’ Organization at Children’s (PPOC), a pediatric primary care network affiliated with Boston Children’s Hospital. The study was approved by the Institutional Review Boards of Harvard University and Boston Children’s. Language model prompts and codebase are shared at: https://github.com/seouri/sprout.

### 2.1. Dataset creation

#### 2.1.1. Cohort selection

The sprout LLM-based diagnostic reasoning engine described here was developed and validated using a large, electronic health records (EHR) dataset of pediatric primary care patients in Massachusetts. Exclusion criteria were applied to yield a cohort of patients with sufficiently long observation periods and to exclude patients with rare clinical events (defined as fewer than 10 occurrences in the database) that may allow for re-identification (Figure 1). From these patients, a de-identified dataset was created using the following discrete EHR data elements as of December 31, 2024: demographics including age, race, and ethnicity; anthropometric measurements such as height, weight, and head circumference; diagnosis codes; specialist referrals; medication and prescription data; laboratory test orders and results; and clinical encounter metadata.

**Figure 1.**
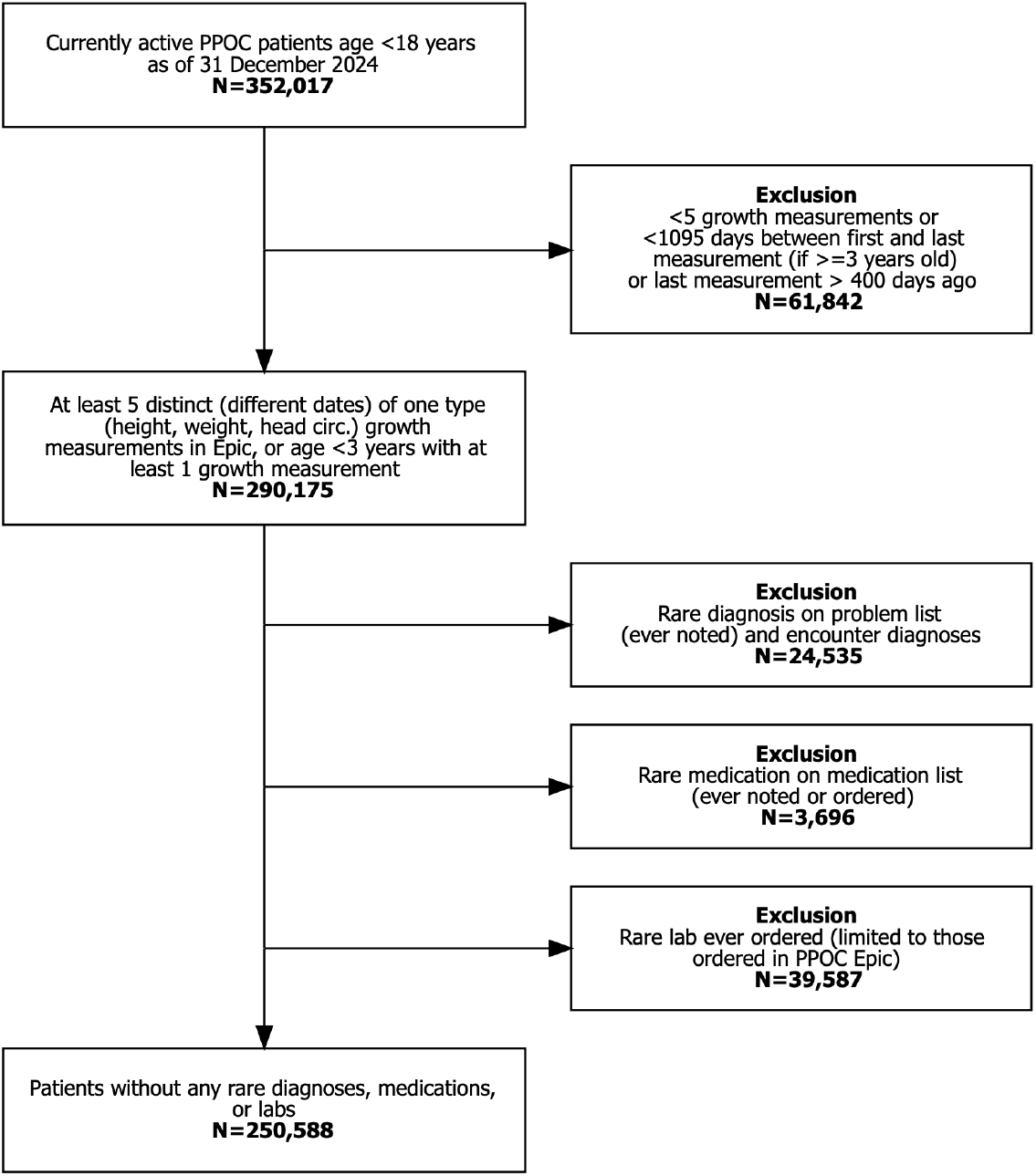
Waterfall diagram detailing cohort inclusion and exclusion criteria.

**Figure 2.**
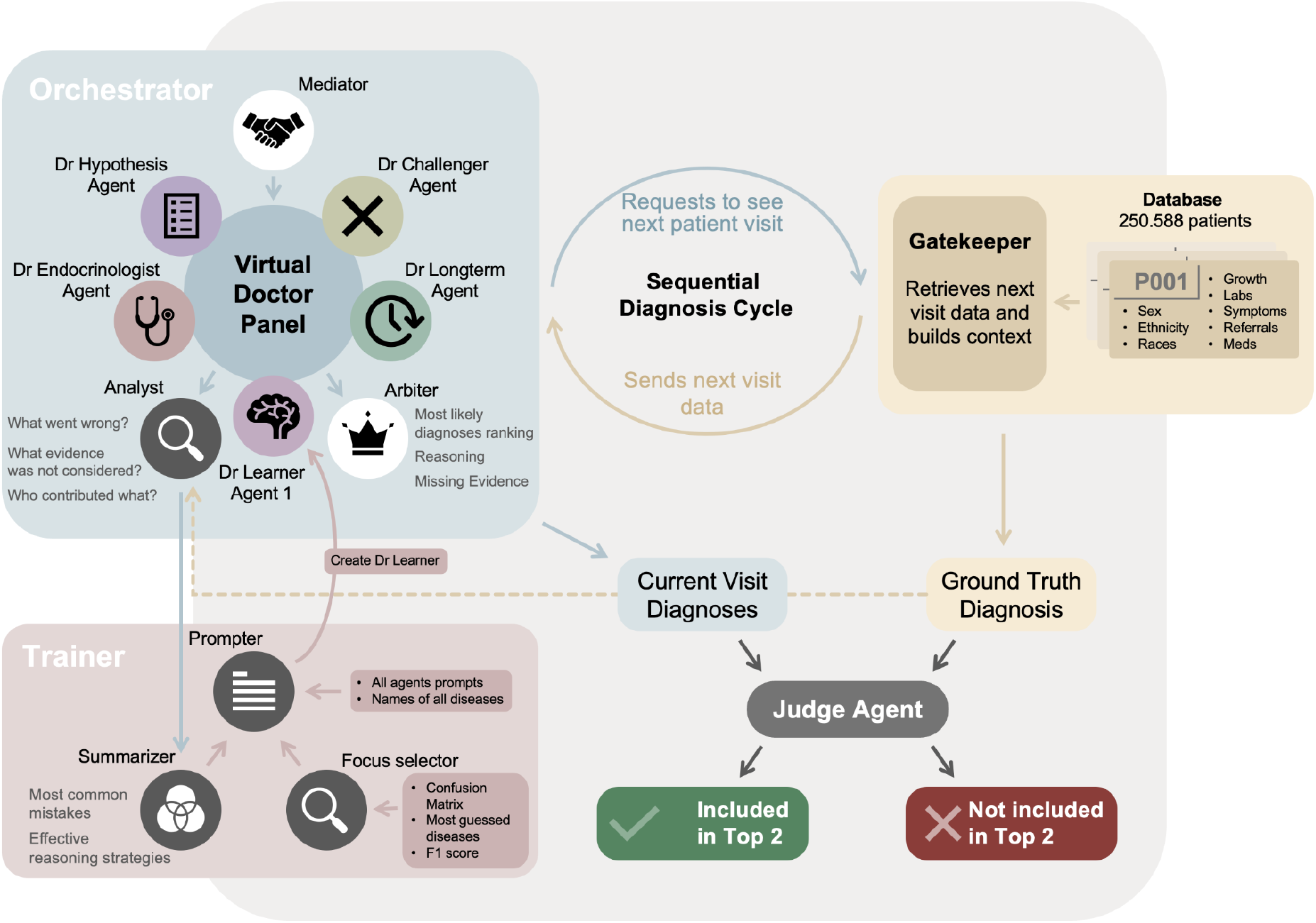
Diagram illustrating the design and data flow for the sprout multi-agent system diagnostic reasoning engine and learning module. The Gatekeeper prepares patient data one visit at a time. This chronological data stream simulates a prospective validation. The Orchestrator module comprises an expert panel that receives the patient data and outputs a differential diagnosis ranked list. The Trainer module reviews misclassifications on the training dataset and uses that to seed a Learner agent for the expert panel. The bottom left denotes how a true positive is defined, if the ground truth diagnosis appears in the top two items of the differential diagnosis ranked list.

#### 2.1.2. Annotations

A subset of growth-related chronic conditions of interest was identified as the initial focus for this study. These target conditions were elaborated through subsequent consensus meetings between clinical subject matter experts consisting of two pediatric endocrinologists and one general pediatrician (Table 1). Ground truth labels were assigned using ICD-10 codes documented in the EHR. The first instance of a diagnosis of interest is recorded as the index date for that condition. Those patients with fewer than 5 visits occurring over at least 3 years before the index date were further excluded. Remaining cases were used to develop a multi-agent diagnostic reasoning engine and test its performance in early diagnosis of growth-related conditions in an experiment detailed below. Patients were assigned to test/train cohorts using an approximately 50-50 split.

**Table 1.** Overview of target diagnoses and patient counts in dataset.

| Disease group | ICD-10 code(s) | Original dataset | Included for training and testing | Assigned to test cohort | Assigned to train cohort |
| --- | --- | --- | --- | --- | --- |
| Hypothyroidism | E03.9 | 271 | 153 | 77 | 76 |
| Type 1 diabetes mellitus | E10 | 464 | 305 | 153 | 152 |
| Pituitary disorders | E23.0, E23.6, E23.7, E34.3 | 566 | 389 | 196 | 193 |
| Crohn’s disease | K50 | 99 | 89 | 45 | 44 |
| Ulcerative colitis | K51 | 57 | 43 | 22 | 21 |
| Celiac disease | K90.0 | 835 | 500 | 250 | 250 |
| Chronic kidney disease | N18 | 59 | 54 | 27 | 27 |
| Genetic syndromes (Marfan’s and Turner’s) | Q96, Q87.4 | 48 | 18 | 10 | 8 |

Controls were designed to approximate a healthy cohort using diagnosis-code-based exclusion criteria. We mapped all diagnosis codes to the Chronic Condition Indicator Refined (CCIR, version 2025.1, HCUP) and retained only patients whose diagnoses are classified as “Not chronic (0)” or “No Determination (9)”.^12^ To reduce the influence of implausible anthropometric entries, we excluded patients with any height or weight Z-score outside [−3.09, 3.09] (0.1 percentile threshold under CDC growth charts).^13^ This procedure yielded 40,452 candidate controls. To align the temporal structure of controls with cases, we sampled a random cutoff time per control to match the age distribution of the diseased cohort and then retained only controls with at least five visits before that cutoff.

One limitation of using the first occurrence of a diagnosis in the EHR as the index date is that a clinician may suspect a particular diagnosis before it is entered in the chart. Therefore, the expert panel curated a gold-standard dataset of 125 patients with 90 (72%) controls and 35 (28%) cases. Each patient was manually reviewed to confirm the ground truth diagnosis and index date corresponding to the first date on which the diagnosis was suspected. In some cases, specialty referral or laboratory testing indicated earlier clinical suspicion and the index date was adjusted accordingly.

### 2.2. Clinical reasoning engine design

We designed sprout to have two physician-facing decision support components that can be integrated into EHR workflows. The first is a binary screening model that flags whether a longitudinal record suggests a possible undiagnosed growth-related condition. A second model outputs a ranked differential diagnosis over a fixed set of disease groups. Depending on the ultimate application, these tools may be implemented in series. The first component is a prompted frontier model. The second component is built from an orchestrated multi-agent harness. Of note, in addition to the raw, structured EHR data, we also feed anthropometric percentiles, z-scores, and growth velocities as inputs to the model. In development, we found the inclusion of these additional abstractions significantly improved system performance.

#### 2.2.1. Screener model

The sprout screener model uses an LLM (GPT-5) prompt to identify children with possible undiagnosed growth disorders while minimizing false alarms. It is prompted with eight clinician-labeled few shot examples taken from the gold-standard dataset, including three positive cases and five controls. Each example includes a short sentence drafted by the expert clinical panel describing the clinical reasoning to guide the decision toward longitudinal patterns rather than isolated measurements. The few-shot samples were excluded from subsequent screener evaluation, yielding 117 test patients with 85 controls and 32 cases. For each patient record, the screener model produces a final boolean output indicating whether the patient should be flagged for further examination.

#### 2.2.2. Multi-agent system diagnostic reasoning engine

The sprout diagnostic reasoning engine comprises two modules: the Orchestrator and Trainer. The Orchestrator module receives structured EHR data and outputs a ranked list of differential diagnoses. The Trainer module is engaged when running the system on the training set. It observes misclassifications and feeds this learning back to the Orchestrator in the form of a prompt for a “learner agent” that participates in the Orchestrator’s multi-agent panel. The Orchestrator is fed EHR data one visit at a time for each patient. This chronological data stream simulates a prospective validation and is mediated by a Gatekeeper agent, which prepares and restricts the input to data available at each visit accordingly. Prompts for each agent are published in the project repository.

The Orchestrator module consists of a panel of language model agents (GPT-4.1 unless otherwise noted) that work in concert to produce a ranked list of differential diagnoses. The panel includes a mediator, expert panel participants, and an arbiter, which synthesizes the panel members’ contributions into a final ranked list. This multi-agent system approach is based on the work by Nori *et al*. at Microsoft.^14^ The Orchestrator module is composed of the following agents:

- **Mediator**: Guides discussion and controls the loop structure. At each step, the Mediator provides the list of allowable diseases the experts must choose from, flags possible measurement errors in the EHR, and provides disease prevalence to calibrate discussions.
- **Dr Hypothesis**: Pediatrician agent that generates and refines hypotheses from longitudinal records, aiming to cover plausible conditions in the allowed disease set. Retrieval-Augmented Generation (RAG) corpus includes “Disorders of growth” by Cabrera.^15^
- **Dr Endocrinologist**: Serves as an endocrinology-focused agent grounded in relevant literature. Interprets growth trajectories and endocrine-related evidence in the structured record and contrasts endocrine explanations with alternative disease groups. RAG corpus includes “Disorders of growth and stature” review article by Braun and Marino.^16^
- **Dr Longterm**: Pediatrician agent focused on long-term patterns and early-stage disease presentations that may only become clear across multiple visits.
- **Dr Challenger**: Challenges the panel to avoid early narrowing of the differential, points out contradictions in reasoning, and highlights missing evidence and alternative explanations.
- **Arbiter**: Decision agent that uses the experts’ current-visit opinions to produce the ranked list of most likely diseases for the visit. Selects diagnoses that are most strongly supported across the panel.
- **Dr Learner**: This is an agent whose prompt is created by the Trainer module. This agent provides meta-knowledge about common misclassifications and recurring reasoning issues observed in prior training runs, with the goal of correcting systematic errors without changing the core roles of other experts.
- **Analyst**: Observes the panel discussion, and during training, is provided the ground truth diagnosis. It creates a structured report (in JSON) of the discussion for each patient and flags recurring reasoning problems and incorrect classifications. This report is provided to the Trainer module.

The Trainer module comprises three agents that iteratively improve the Orchestrator module by adding metaknowledge about recurring errors. The Trainer module was run on the training set of patients specified in Section 2.1.2. The resulting guidance is encoded in a prompt for the Learner agent that participates in the Orchestrator module expert panel discussion.

We also experimented with rewriting the prompts of all Orchestrator experts. While this occasionally improved performance, it more often led to over-prediction of the weakest class from the training dataset because the experts received similar updated instructions, even when the Prompter model was instructed to keep expert roles distinct. In contrast, adding a single Learner agent was more stable because it introduced a focused perspective without changing the behavior of the existing experts. Adding a second Learner did not yield significant gains in performance. The agents in the Trainer module are described below:

- **Summarizer**: Aggregates the patient-level JSON report from the Analyst agent and compiles recurring themes around classification errors. An example output is included as a supplement on the project repository.
- **Focus selector**: Calculates a confusion matrix, F1-scores, and classification distributions from the training run and submits them to the Prompter for additional context on how to improve future performance.
- **Prompter (Claude Opus 4.5)**: Generates the Learner agent prompt using the outputs from the Summarizer and Focus selector.

### 2.3. Experimental design

We present the findings of three experiments: two screening experiments and one diagnostic experiment. Screening model performance was assessed with two experiments: first on the 117 test patients in the gold-standard dataset and on a randomly selected 300-patient cohort of controls who are either presumably healthy or undiagnosed. The third experiment evaluating the performance of the diagnostic reasoning engine ran the multi-agent system on all disease cases in the withheld testing dataset. Data was fed to the sprout system one visit at a time leading up to the eventual date of diagnosis in order to assess the system’s capability for early disease detection.

#### 2.3.1. Screening experimental design

The first experiment uses the 117 patients from the manually reviewed gold-standard dataset, calculating sensitivity, specificity, and 2 × 2 confusion matrices both one year before and on the day of the index date (date of diagnosis for disease cases or the random cutoff point for controls).

In a second experiment, 300 patients were randomly selected from the control cohort. These are patients who are either healthy or undiagnosed. The cases were presented to the sprout screener model using all available longitudinal clinical data. Cases that were positively identified by the screener were presented to the expert panel of three clinicians for manual review to confirm whether the prediction was appropriate. Each expert reviewed cases independently and voted whether there was a high suspicion for an underlying growth-related disorder. A positive prediction by sprout was deemed a true positive if all three clinicians agreed there was a high likelihood of undiagnosed condition. Otherwise, it was considered a false positive.

#### 2.3.2. Multi-agent system diagnostic reasoning engine experimental design

For each patient in the disease cohort, structured, longitudinal EHR data is fed to the multi-agent system one visit at a time from two years before diagnosis through the diagnosis date. An initial run was performed on the training set to furnish the Learner agent prompt.

After a training run, the system was then applied to the withheld test set of patients to calculate its performance. For each disease group, sensitivity (the proportion of positive cases correctly identified by the multi-agent system) was calculated at each time relative to the date of diagnosis. A model output was defined as a true positive if it included the ground-truth diagnosis in the top two diagnoses in the differential diagnosis ranked list.

## 3. Results

Table 2 presents the confusion matrices summarizing the performance of the screener model at discriminating between disease and healthy patients at two time points: one year before the index date (date of diagnosis or random cutoff point for controls) and day of index date.

**Table 2.** Screening performance identifying patients with and without growth conditions 1 year before (sensitivity 28%, specificity 98%) and day of index date (sensitivity 47%, specificity 100%)

|  |  | Prediction 1 year before |  | Prediction on index date |  |
| --- | --- | --- | --- | --- | --- |
|  |  | Positive screen | Negative screen | Positive screen | Negative screen |
| Ground truth | Growth disorder | 9 | 23 | 15 | 17 |
|  | Healthy | 2 | 83 | 0 | 85 |

Of the 300 control cohort patients, the screener model identified 15 as potentially having a growth-related condition. Of those, the expert panel unanimously voted that five were clinically concerning, representing a positive predictive value of 5/15 (33%). Two of those cases are discussed below for illustrative purposes (Figure 3).

**Figure 3.**
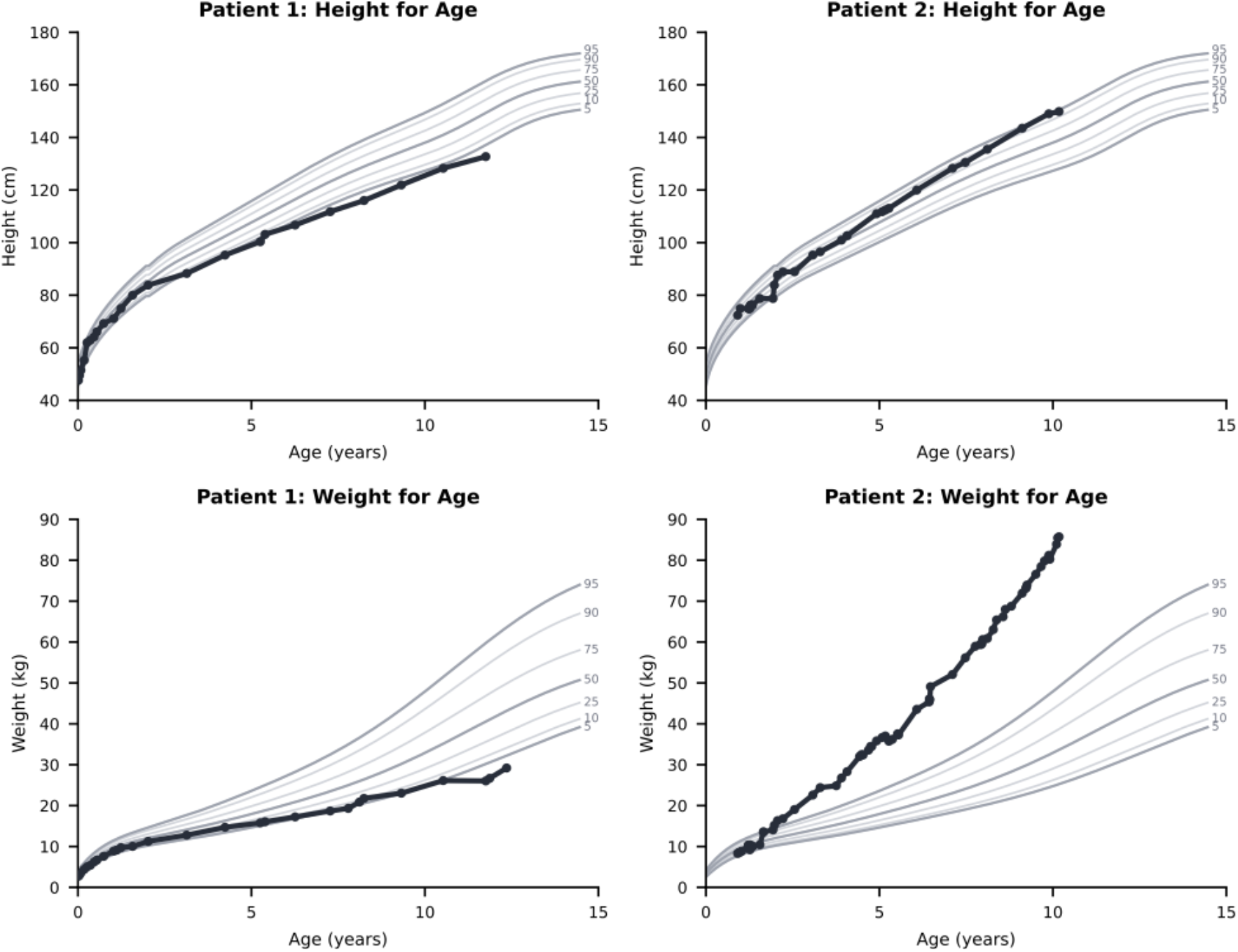
Growth charts for undiagnosed patients identified by the screener model. (Left) concern for celiac disease or other enteropathy and (Right) concern for monogenic obesity.

The first case demonstrates consistent deceleration in height velocity by age 3 that worsens into late childhood, concomitant with suboptimal weight gain. All panel members flagged this concerning pattern as clinically significant, a pattern consistent with enteropathies such as celiac disease.

The second case demonstrates severe, early-onset weight gain with the absence of the typical childhood body mass index nadir, preceding accelerated height velocity. This pattern is characteristic of monogenic obesity. In neither case were there relevant diagnoses, specialist referrals, or laboratory testing (e.g., antibody testing for celiac or genetic testing for obesity).

Figure 4 demonstrates the sprout diagnostic reasoning engine’s ability to diagnose specific conditions earlier than standard of care. Performance is variable across conditions of interest. At one extreme, the diagnostic system is able to identify patients with type 1 diabetes mellitus 1 year before diagnosis with a sensitivity of 81%. At the other extreme, sensitivity for identifying patients with Crohn’s disease 1 year before diagnosis is only 20%. In the middle, diagnostic sensitivity 1 year before diagnosis is 56% for pituitary disorders and 44% for celiac disease.

**Figure 4.**
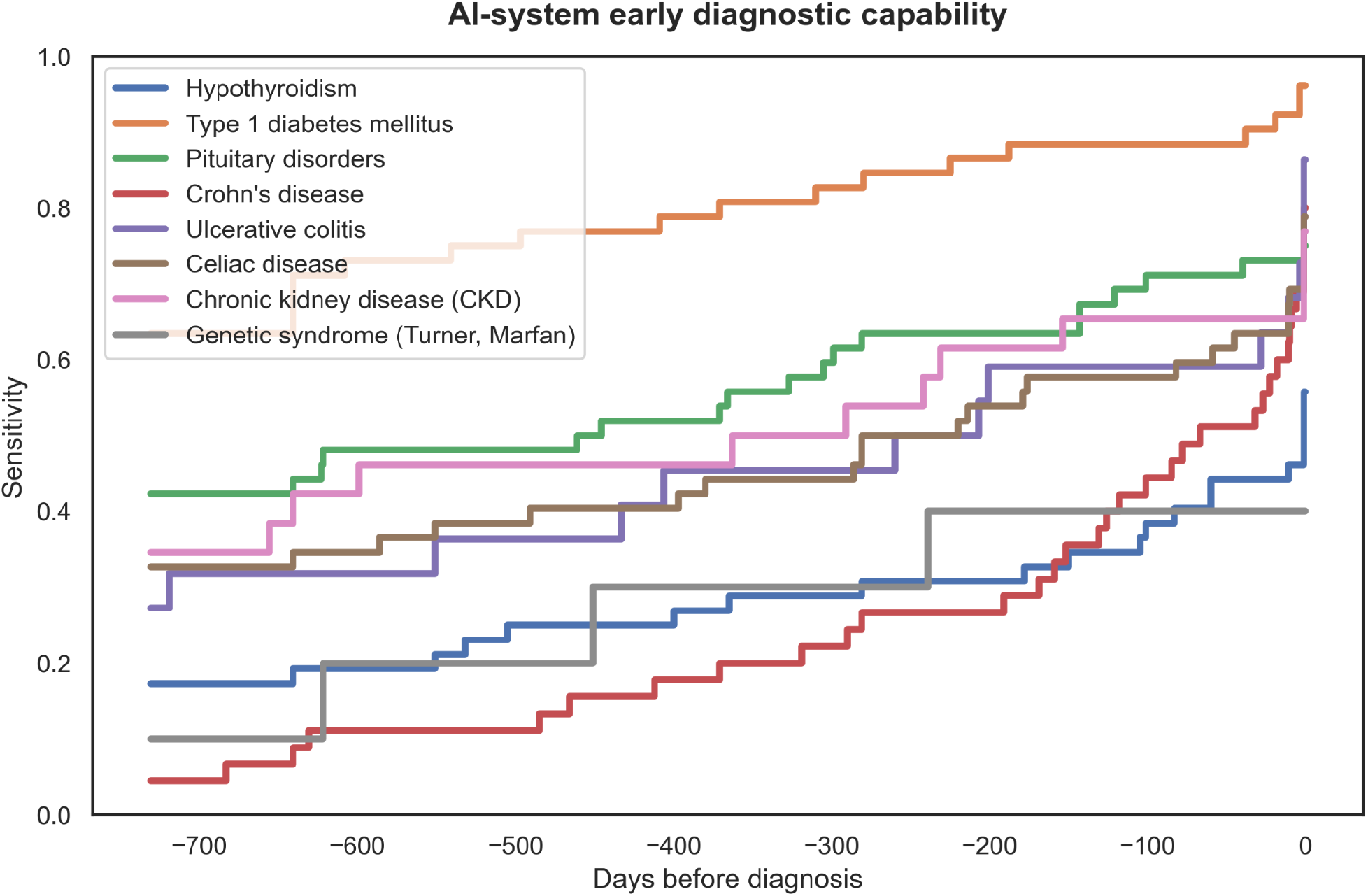
sprout diagnostic reasoning engine’s sensitivity as a function of days prior to documented diagnosis. Each trace represents a distinct condition evaluated during the walk-forward simulation. Sensitivity is defined as the proportion of patients for whom the system successfully placed the ground-truth condition within the top two of the ranked diagnosis list. Sensitivity across multiple conditions at time points months to years prior to the index date demonstrates the system’s ability to recognize latent clinical patterns and its potential to reduce diagnostic delays.

## 4. Discussion

In this study, we developed and retrospectively validated SPROUT, a generalized, multi-agent LLM system designed to enable early detection of pediatric growth-related conditions. Our results demonstrate the feasibility and clinical utility of applying LLMs to the diagnosis of growth-related pediatric conditions. The sprout screener model effectively identified underlying pathology in an undiagnosed cohort with a high degree of clinical relevance upon expert review (33% PPV).

Furthermore, when evaluating the diagnostic reasoning engine using a walk-forward simulation, the system successfully identified conditions like type 1 diabetes mellitus up to one year before the actual clinical diagnosis with 81% sensitivity, and celiac disease with 44% sensitivity.

An important finding of our screening experiment is the sprout performance profile, which intentionally favors specificity. On the gold-standard dataset, the screener achieved 98% specificity 1 year from the index date and 100% specificity on the index date at the expense of lower sensitivity, 28% and 47% respectively. In the context of primary care, this high-specificity profile is critical. While a lower sensitivity means the system will not catch every latent case, the ability to accurately flag a substantial fraction of these conditions months or years earlier while minimizing false alarm burden represents an improvement over the current standard of care, where diagnostic delays lead to avoidable morbidity. When considering the spectrum of growth-related pathologies, a delayed diagnosis may entail temporary decreases in quality of life (e.g., acquired hypothyroidism) or persistent morbidity (e.g., late diagnosis of an intracranial tumor).

sprout presents a novel clinical application of LLMs in pediatrics, addressing the problem of diagnostic delay in pediatric growth disorders. Furthermore, this study advances the technical approach of applying LLMs in pediatric practice by moving beyond single-prompt models to a learning, multi-agent orchestrated panel.^17–19^ Importantly, we introduce a training mechanism via the Learner agent. Standard approaches to updating LLM behavior can be problematic. Fine-tuning is computationally expensive, while modifying the base prompts of existing agents resulted in over-prediction of minority classes and blurring of distinct agent roles. Synthesizing misclassifications into a dedicated Learner agent enabled effective iterative model improvement.

Furthermore, the experimental design of streaming longitudinal patient records, as opposed to validating on static vignettes, also better simulates the system’s ability to deploy prospectively in a primary care setting. An additional finding of interest is that in our experimentation, the frontier models did not appear to automatically generate abstractions of percentiles and velocities from the raw anthropometric data. In order to improve performance, these values were calculated and explicitly fed as inputs to the model. As models improve, the need for this and other parts of the harness may disappear, but at this moment, we found them necessary.

While Kohane *et al*. established the utility of computational growth trend analysis, their methods largely relied on expert-derived rigid mathematical thresholds.^7^ More recent machine learning approaches have shown promise but are disease-specific. For example, Daniel *et al*. achieved a 71.6% sensitivity for type 1 diabetes at 90 days pre-diagnosis using a SuperLearner model.^9^ sprout achieved 81% sensitivity a full year prior. Similarly, Dreyfuss *et al*. reported strong predictive performance for celiac disease one year out (AUC 0.86).^10^ sprout achieved a 44% sensitivity at the same time point, but did so within a generalized framework. Unlike disease-specific models, our model continuously evaluates a broad differential, making it well suited to primary care, where any single disorder is rare but their combined burden is substantial.

There are several key limitations to acknowledge. First, the validation is retrospective. Second, the multi-agent diagnostic reasoning engine was evaluated exclusively on disease cases; thus, we lack a formal measure of false-positivity for the diagnostic module, and the top-two criterion carries a non-trivial chance baseline rate. These sensitivities are best read as a measure of when the signal of each condition becomes detectable in the structured record, not as deployment-level diagnostic accuracy. Relatedly, the artificial case-control matching design used for the screening experiments is not ideal for accurately estimating positive and negative predictive values, nor does it perfectly estimate real-world impact in an unselected population. Third, the sprout screening and diagnostic reasoning engines were evaluated in isolation rather than in tandem, meaning the cumulative performance of a fully serial, end-to-end pipeline remains unmeasured. Additionally, the reported positive predictive value is measured against unanimous expert suspicion rather than confirmed disease, as these patients are undiagnosed by definition. Fourth, the index date is the first coded instance of a diagnosis and may postdate the clinician’s initial suspicion. Because the input stream includes specialist referrals and laboratory testing, some predictions may reflect a workup already in progress rather than an opportunity for earlier diagnosis.

Fifth, performance was not evaluated across demographic subgroups, across which growth references and practice patterns vary. Subgroup calibration is a prerequisite to deployment. sprout also relies on proprietary models and each patient was evaluated in a single run, so performance may vary with model version and across repeated runs in ways we did not quantify. Finally, a significant limitation is the omission of unstructured clinical notes from the dataset.

Future iterations of this work will directly address these limitations. We envision integrating unstructured clinical notes into the data stream, which we hypothesize will improve sensitivity for symptom-heavy diseases like inflammatory bowel disease. Furthermore, future research must focus on deploying this system in an unselected, prospective clinical cohort to accurately measure real-world predictive values and evaluate physician interaction with the diagnostic alerts.

In conclusion, multi-agent LLM systems equipped with iterative learning mechanisms hold promise for interpreting longitudinal health records. By recognizing subtle, long-term clinical patterns, such tools may reduce diagnostic delay in identifying pediatric growth-related disorders.

## Data Availability

Patient data are not available per institutional policy. Code related to experimental design is made publicly available.

https://github.com/seouri/sprout

## 5. Supplementary Materials

Language model prompts and codebase for computational experiments are publicly available at https://github.com/seouri/sprout

## 6. Acknowledgments

This work was funded by Charles H. Hood Foundation and Yosemite.

## References

1. Committee on Practice and Ambulatory Medicine, Bright Futures Periodicity Schedule Workgroup. 2023 Recommendations for Preventive Pediatric Health Care. Pediatrics. 2023;151(4):e2023061451.

2. Chen RS, Shiffman RN. Assessing growth patterns–routine but sometimes overlooked. Clin Pediatr (Phila). 2000 Feb;39(2):97–102.

3. Singh H, Thomas EJ, Wilson L, Kelly PA, Pietz K, Elkeeb D, et al. Errors of diagnosis in pediatric practice: a multisite survey. Pediatrics. 2010 Jul 1;126(1):70–9.

4. Kappelman MD, Brensinger C, Parlett LE, Hurtado-Lorenzo A, Lewis JD. Prevalence of pediatric inflammatory bowel disease in the United States: Pooled estimates from three administrative claims data sources. Gastroenterology. 2025 May 1;168(5):980–2.e2.

5. Sulkanen E, Repo M, Huhtala H, Hiltunen P, Kurppa K. Impact of diagnostic delay to the clinical presentation and associated factors in pediatric inflammatory bowel disease: a retrospective study. BMC Gastroenterol. 2021 Oct 7;21(1):364.

6. Stochholm K, Juul S, Juel K, Naeraa RW, Gravholt CH. Prevalence, incidence, diagnostic delay, and mortality in Turner syndrome. J Clin Endocrinol Metab. 2006 Oct 1;91(10):3897–902.

7. Haimowitz IJ, Kohane I. Automated trend detection with alternate temporal hypotheses. Int Jt Conf Artif Intell. 1993 Aug 28;146–51.

8. Kohane IS, Haimowitz IJ. Hypothesis-driven data abstraction with trend templates. Proc Annu Symp Comput Appl Med Care. 1993;444–8.

9. Daniel R, Jones H, Gregory JW, Shetty A, Francis N, Paranjothy S, et al. Predicting type 1 diabetes in children using electronic health records in primary care in the UK: development and validation of a machine-learning algorithm. Lancet Digit Health. 2024 Jun 1;6(6):e386–95.

10. Dreyfuss M, Getz B, Lebwohl B, Ramni O, Underberger D, Ber TI, et al. A machine learning tool for early identification of celiac disease autoimmunity. Sci Rep. 2024 Dec 28;14(1):30760.

11. Brodeur PG, Buckley TA, Kanjee Z, Goh E, Ling EB, Jain P, et al. Performance of a large language model on the reasoning tasks of a physician. Science. 2026 Apr 30;392(6797):524–7.

12. Chronic Condition Indicator Refined (CCIR) for ICD-10-CM [Internet]. Available from: https://hcup-us.ahrq.gov/toolssoftware/chronic_icd10/chronic_icd10.jsp. Accessed 20 June 2025.

13. Growth Charts - Percentile Data Files with LMS Values [Internet]. 2024. Available from: https://www.cdc.gov/growthcharts/cdc-data-files.htm. Accessed 20 June 2025.

14. Nori H, Daswani M, Kelly C, Lundberg S, Ribeiro MT, Wilson M, et al. Sequential diagnosis with language models. arXiv:2506.22405. 2025.

15. Cabrera S. Disorders of Growth. Board Review Course in Pediatric Endocrinology; 2025. Available from: https://pedsendo.org/wp-content/uploads/2025/04/Growth_Cabrera_FINAL.pdf. Accessed 20 June 2025.

16. Braun LR, Marino R. Disorders of growth and stature. Pediatr Rev. 2017 Jul 1;38(7):293–304.

17. Huang T, Tse G, Pageler NM, Bannett Y. Large language models using clinical text in pediatrics: A scoping review. JAMA Netw Open. 2026 Mar 2;9(3):e262443.

18. Young CC, Enichen E, Rivera C, Auger CA, Grant N, Rao A, et al. Diagnostic accuracy of a custom large language model on rare pediatric disease case reports. Am J Med Genet A. 2025 Feb;197(2):e63878.

19. Barile J, Margolis A, Cason G, Kim R, Kalash S, Tchaconas A, et al. Diagnostic accuracy of a large language model in pediatric case studies. JAMA Pediatr. 2024 Mar 1;178(3):313–5.

